# Ultrasound-guided skeletal muscle biopsy technique permits measurement of structural, functional, cellular and biochemical properties

**DOI:** 10.64898/2025.12.30.25343237

**Authors:** Addison Barber, Amber Willbanks, Guadalupe Meza, Jeremie L.A. Ferey, Gretchen A. Meyer, Sudarshan Dayanidhi, W. David Arnold, Richard L. Lieber, Ishan Roy

## Abstract

Human muscle biopsies are often required to study or diagnose diseases. However, traditional approaches are challenging due to limited sample size, quality, or patient discomfort. Fine-gauge needle biopsies (≥14-gauge), present an alternative but yield insufficient sample sizes for histology or function. Ultrasound guidance, coupled with vacuum-assisted, single needle-insertion multiple sampling addresses these challenges. In 19 healthy participants (mean age: 30.1±10 years, 42% male), 2-3 samples were collected from a single needle insertion into the vastus lateralis (VL) and tibialis anterior (TA). Summed VL and TA sample masses averaged 148±38mg and 166±64mg, with dimensions of 15.83±8 x 2.9±0.6mm^2^ (VL) and 15.07±7 x 3.1±0.9mm^2^ (TA). VL had a mean fiber cross-sectional area of 4,347±1,931µm^2^, with 221±86 fibers quantified. Samples were of sufficient size and quality for thorough analyses from a single biopsy procedure, including mitochondrial respirometry, RT-PCR, collagen content, and biomechanical function. Fibers produced typical isometric stress values of 187kPa with a passive modulus of 239kPa (peak) and 79kPa (stress-relaxed). This procedure was well tolerated, with an average immediate pain rating of 1.5±1 (range:0-4, scale: 1-10) and 24-hour follow-up rating of 1.7±1 (range:0-4). This report describes an approach that yields high-quality muscle samples suitable for histological and biochemical analyses while minimizing discomfort.

## Introduction

The muscle biopsy has long been a cornerstone in the diagnosis and study of human neuromuscular and systemic diseases, enabling direct analysis of muscle structure. Traditional methods, including the Bergström needle, open biopsy, and, more recently, microbiopsy, have provided valuable insights but suffer from limitations such as small sample sizes, procedural variability, and poor tissue quality [1]. These shortcomings hinder reproducibility and restrict the scope of robust downstream analyses, particularly those that multiple involve physiological, molecular, or histological studies. Given the fundamental information gleaned from human muscle tissue in clinical and research settings, we believed it was imperative to address these limitations.

Here we present a protocol that uses a vacuum assisted soft tissue biopsy device (BD EleVation Breast Biopsy Device, hereto referred to as the “biopsy device”) combined with handheld ultrasound guidance. The cordless, lithium battery powered biopsy device has Single-Insertion-Multiple-Samples (SIMS) technology that allows collection of multiple muscle samples with a single needle insertion. Ultrasound guidance allows orientation of the biopsy needle parallel with muscle fibers, resulting in samples containing relatively long muscle fiber bundles. Ultrasound also allows direct visualization of vasculature and nerves, that can be avoided, preventing adverse events. Overall, this approach offers several key advantages over previous methods, including larger and more consistent sample size that allows simultaneous histological, biomechanical, and biochemical analysis of muscle samples, reduced patient discomfort and risk, and expanded access to muscle tissue beyond the operating room.

### Applications of the method

The muscle biopsy plays a crucial role in diagnosis of muscular dystrophies, congenital myopathies, and other neuromuscular disorders, often revealing characteristic pathological changes [2]. Additionally, skeletal muscle may exhibit unexpected histological or biochemical alterations in a range of acute and chronic conditions, even in the absence of overt clinical signs, making muscle pathology a valuable tool enabling both differential diagnosis and research into disease mechanisms and progression [3]. These pathological changes contribute to the functional decline and muscle weakness, associated with aging [4–6] and diseases such as cancer [7] and chronic heart failure [8,9], arising not only from loss of muscle mass but also to deterioration of intrinsic muscle-fiber properties. Mechanical testing of muscle fibers represents a powerful tool for studying muscle function at the cellular level [10], but its use in human research has been restricted by limitations in biopsy sample size and fiber integrity.

This improved biopsy technique is suitable for a range of clinical and research purposes and is distinctive in its ability to yield large, high-quality samples that can be distributed to multiple routes of downstream histological, biomechanical, and biochemical analysis from the same muscle sample (Fig. 1). Its ability to yield intact, well-preserved tissue samples also facilitates the study of muscle fiber type distribution, fiber cross-sectional area (fCSA), muscle fiber mechanics, and molecular markers indicative of disease progression or therapeutic response.

**Figure 1:**
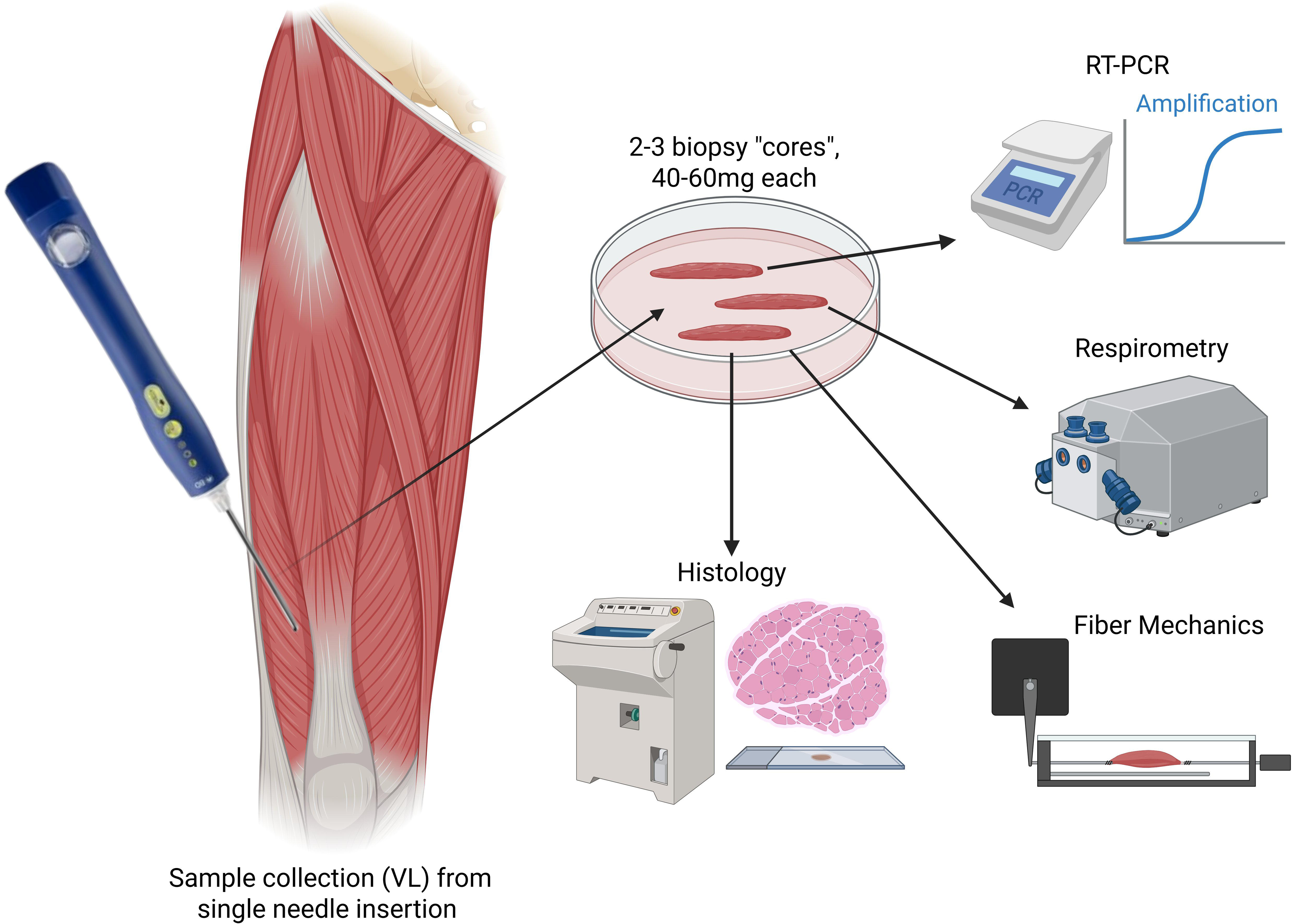
Schematic representation of a VL biopsy depicting biopsy device, yield, and interchangeable sample allocation for histological, mechanical, and metabolic analysis.

### Comparison with other methods

Traditional muscle biopsy methods, while foundational, exhibit several critical limitations that hinder their broad application in both research and clinical settings. For instance, the Bergström needle biopsy can yield inconsistent sample size and requires multiple insertions for multiple samples, which is then compounded by the large needle gauge (6G), that causes significant patient discomfort [11]. Alternatively, several microbiopsy or fine-gauge needle (>16G) biopsy devices exist but are severely limited by low sample mass.

While these samples can be used in physiological analysis of muscle mechanics [12], the small samples restrict the potential for additional and comprehensive biochemical analysis or may fail to capture representative muscle structure or fiber type distribution [1,13]. The open biopsy, on the other hand, provides a much larger sample but is significantly more invasive by requiring a >1cm incision through the skin, subcutaneous tissue, and muscle fascia, resulting in visible scaring, higher risk of infection, and extended recovery times [1,14]. Due to these complications, the open biopsy is typically restricted to the surgical setting, limiting the potential population to surgical candidates. Finally, simple Bergström and microbiopsy methods lack the precision necessary to target specific regions within a muscle or to consistently avoid vascular structures without the addition of ultrasound guidance.

The technique described in this report addresses these shortcomings. By integrating real-time ultrasound guidance, this method allows precise visualization of the biopsy site, including the ability to target regions of abnormal or disordered muscle. Ultrasound guidance also ensures consistent alignment with muscle fibers and avoids major blood vessels thus reducing procedural risks and sample quality variability. Moreover, SIMS technology, with its internal vacuum mechanism and sample collection receptacle, permits collection of multiple samples through a single needle insertion, enabling comprehensive analyses without compromising patient comfort or tissue preservation. Collectively, these innovations represent a superior alternative to traditional methods, offering the combination of safety, efficiency, and insight into physiology and function. While ultrasound guidance and self-contained vacuum biopsy systems have been reported previously [15–18], the technical detail necessary to reproduce the use of these integrated technologies is still lacking. Thus, this article describes ultrasound guided vacuum biopsy in detail to allow readers to implement this technique for clinical or research use.

### Protocol and validation studies

Pilot experiments were initially conducted in postmortem rabbit quadriceps muscles to assess sample yield. Subsequent preliminary cadaveric human studies yielded critical insights to inform and develop our biopsy procedure in the VL and TA particularly in addressing anatomical variation among humans. This study was deemed IRB exempt by our institutional IRB (STU00220158). These preclinical studies also allowed us to refine the procedural timing, and sample processing after collection.

In the current study of 19 healthy subjects (mean age: 30.1 years±10.6 (SD), 42% male), we collected 2-3 muscle samples from a single needle insertion with the biopsy device from vastus lateralis (VL) and tibialis anterior (TA) muscles.

### Procedure design

#### Expertise needed

An individual trained in ultrasound guided procedures is required to perform the biopsy in a sterile field and to monitor the participant for signs of distress (hereafter referred to as the “sterile operator”). A research assistant (RA) or technician is needed to handle non-sterile equipment and process the muscle sample(s) in a timely manner.

#### Setting /space

Patients should lay supine in a hospital bed. A procedure space with curtains or an isolated room is preferred to ensure sterility, patient privacy and comfort. Ample space bedside is required for the physician and team to operate; we suggest minimum 1.5m^2^. Separate workspaces are required for sterile procedures and nonsterile specimen processing.

### Considerations before the muscle biopsy

Accurate site selection is critical to the success of the biopsy procedure. The choice of muscle and specific biopsy site is influenced by factors such as the research objective, anatomical differences among muscles, and patient-specific considerations. The VL is frequently chosen due to its accessibility, size and well-characterized fiber type distribution [19]. For pathological conditions hallmarked by proximal weakness, the VL provides valuable insights without being overly affected by disease-related replacement of muscle tissue with adipose or connective tissue [2]. The TA, on the other hand, is often selected for studies that focus on distal muscle function as it plays a key role in ankle dorsiflexion [20,21]. Its superficial location and functional relevance to everyday movement make it ideal for examining disease-specific changes or therapeutic responses [22]. These two muscles were chosen to optimize the proposed protocol, though ultrasound guidance allows future users to adapt this protocol safely to other muscle groups.

It is also important to consider potential sources of error that may interfere with sample collection and results; a few of these nuances are reported here. Placement or angular rotation of the leg may differ among muscle biopsy sites and should be consistent between participants.

Muscle activation may be elicited during biopsy collection which may create small movements of the biopsy device. Finally, the movement and mechanical sounds of the biopsy device create shadows or feedback interference with the ultrasound and should be expected.

## Materials

### Human Subjects

Participants aged 18+ years gave written informed consent to undergo the muscle biopsy procedure approved by the Northwestern University Institutional Review Board on 6/14/2024 (IRB ID: STU00221110). All methods were performed in accordance with the relevant guidelines and regulations. Permission was obtained to publish all images information/images in an online open access publication.

### Sterile equipment

A full list of sterile equipment required by the sterile operator is listed in Table 1 and pictured in Fig. 2c.

**Figure 2:**
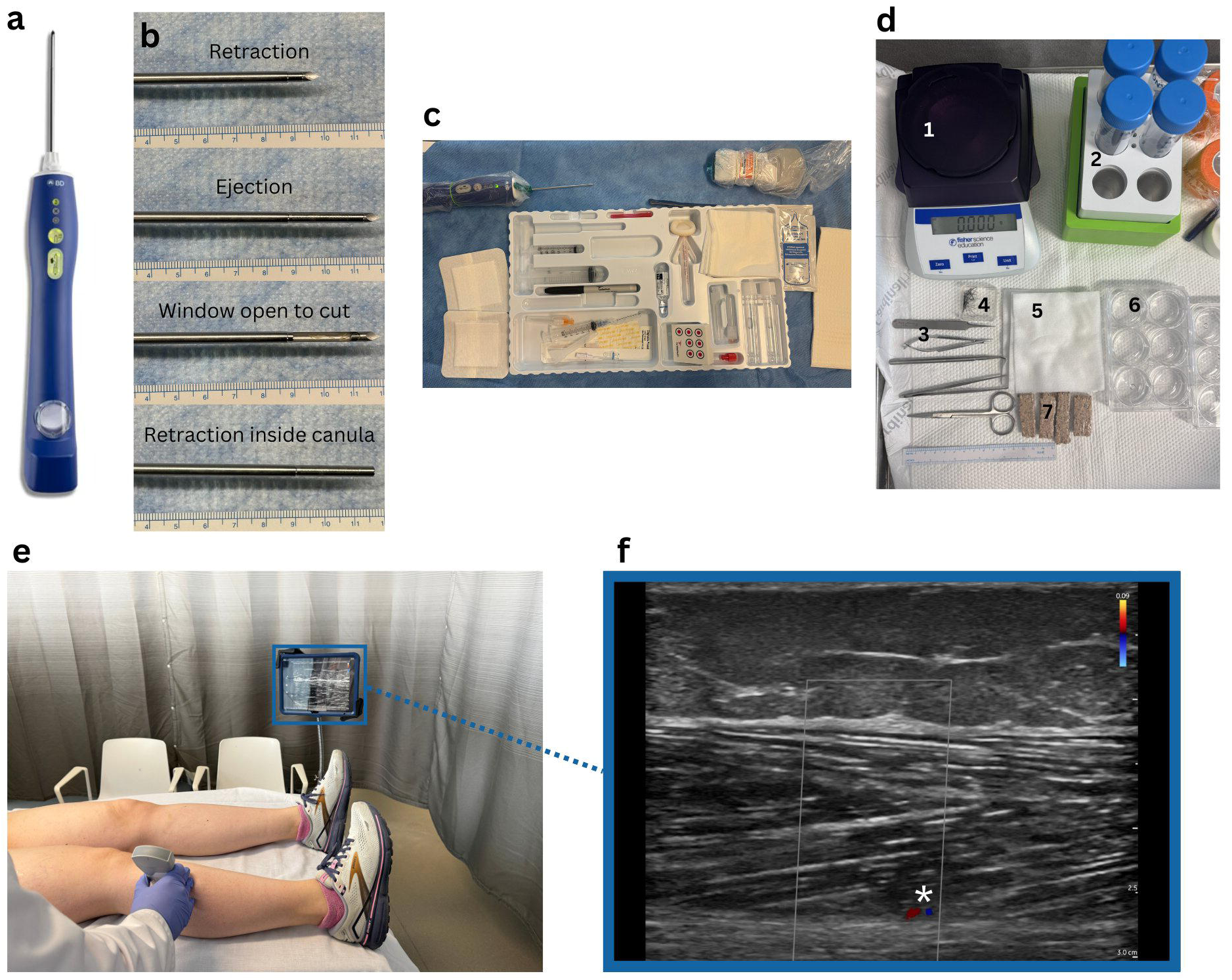
Muscle biopsy preparation. A) BD EleVation Breast Biopsy Probe. B) Stages of needle movement for sample collection: needle retracted, needle ejected, biopsy window open to cut samples, needle retracted inside canula for collection. C) Sterile procedure setup with biopsy device and ultrasound sterilized alongside soft tissue biopsy tray. D) Non-sterile research setup with (1) scale, (2) 50ml tubes in pre-cooled rack, (3) tools for sample handling including fine tip forceps, needle drivers, serrated tip forceps, scissors, and fine tip probe, (4) minute stainless steel pins, (5) gauze to remove blood/debris, (6) 6-well plate with 1x PBS for successive washing of samples, and (7) cork wrapped in parafilm for sample storage. E) Nonsterile ultrasound visualization of biopsy site in the Tibialis Anterior. F) Image from ultrasound application with Doppler effect on. * indicates and identified region of blood flow to be avoided during sample collection.

**Table 1:**
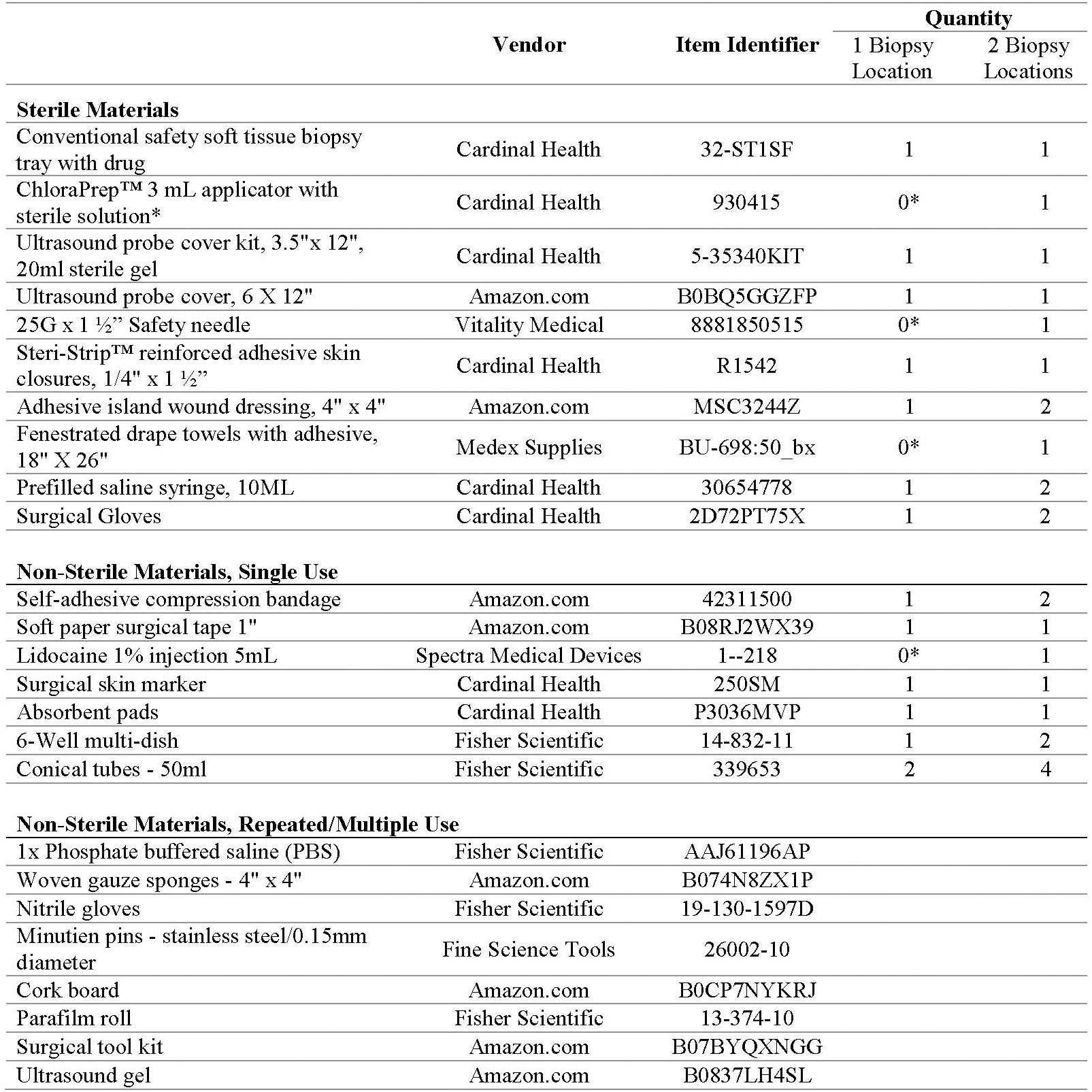
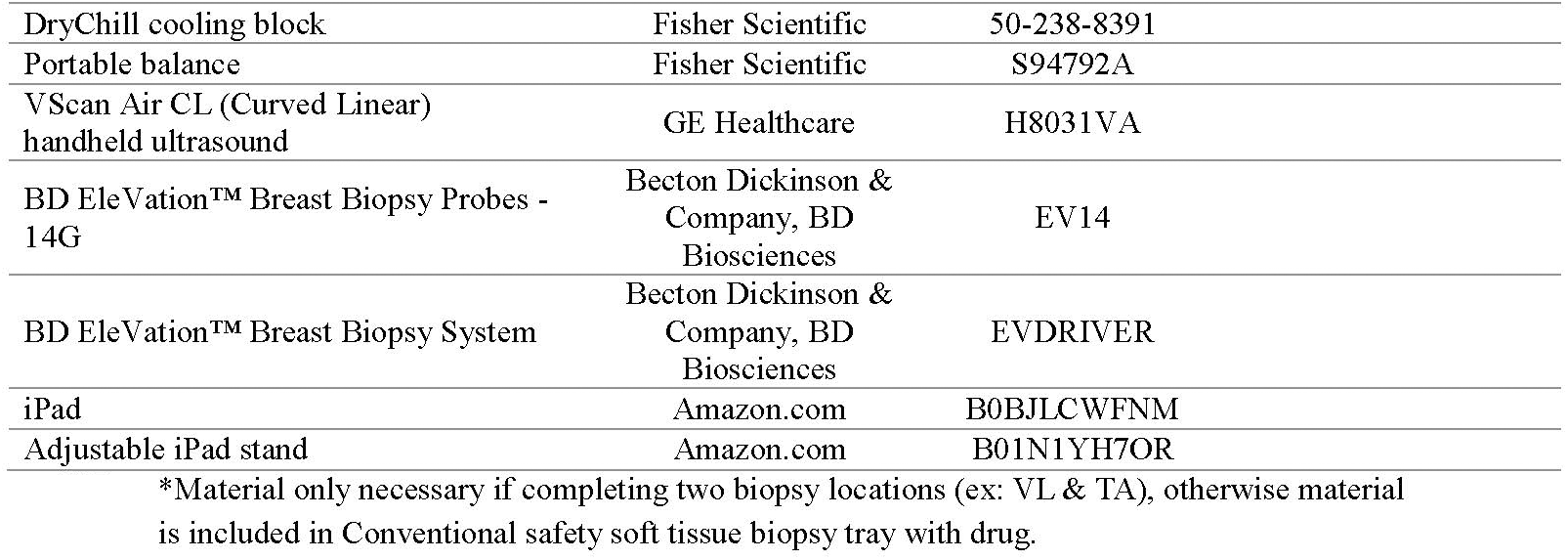
Materials, vendors, and item identifiers of all materials used for biopsy procedure and immediate specimen processing.

### Non-sterile equipment

Non-sterile equipment is necessary to process and store specimens handled by the RA in a non-sterile field located at the periphery of the sterile field. A full list of non-sterile equipment is listed in Table 1 and pictured in Fig. 2d.

### Overview of phases of procedure

*Participant screening:* Aspects of a participant’s medical history were recorded to assess inclusion and exclusion/contraindication criteria. Information such as current medications, including any anti-coagulants, the conditions under which the anti-coagulation may be withheld, any bleeding or clotting disorder, history of chronic infection, and any other significant current or past medical diagnosis. In the presence of any of these factors, the operator performing the biopsy procedure should consult with a physician to determine if the muscle biopsy is contraindicated, as these cases may have a higher risk of complication.

Information about past needle exposures or procedures is also helpful for the research team to anticipate how the participant will tolerate the procedure. The participant should be informed that adverse reactions such as lightheadedness during past needle exposure increase the likelihood of lightheadedness, dizziness, or vasovagal reaction during the biopsy.

For the purposes of our study, exclusion criteria were <18 years of age or anyone unable to give informed consent, the risk, presence, or history of cancer, neurological, or muscular disease, and any contraindication for a muscle biopsy as detailed above.

#### Selection and marking of biopsy site

The VL and TA are superficial lower limb muscles that are relatively easy to access in a clinical setting and are relevant to normal physical function and activities of daily living. Initial non-sterile, ultrasound visualization is critical to differentiate target muscles from nearby anatomical structures, determine the initial angle of needle entry (to acquire samples parallel to fascicles), and identify vasculature and nerves to be avoided during the biopsy (Fig. 2e,f). Both biopsy locations, and ultrasound probe placement sites, should be marked with a skin-safe pen before the sterile field is established.

#### Preparation of the skin, lidocaine

The sterile operator cleans and sterilizes the skin. Then, lidocaine is injected into the sub-dermal layer of the intended needle insertion site (Fig. 3a). This generally represents the most painful phase for the participant. After 60 seconds (to allow lidocaine to act), an additional anesthetic is then injected into deeper subcutaneous and through fascial plane, following the predicted path of the biopsy needle. The depth of subcutaneous tissue should determine the degree and depth of needle insertion to ensure adequate lidocaine delivery to the muscle fascia.

**Figure 3:**
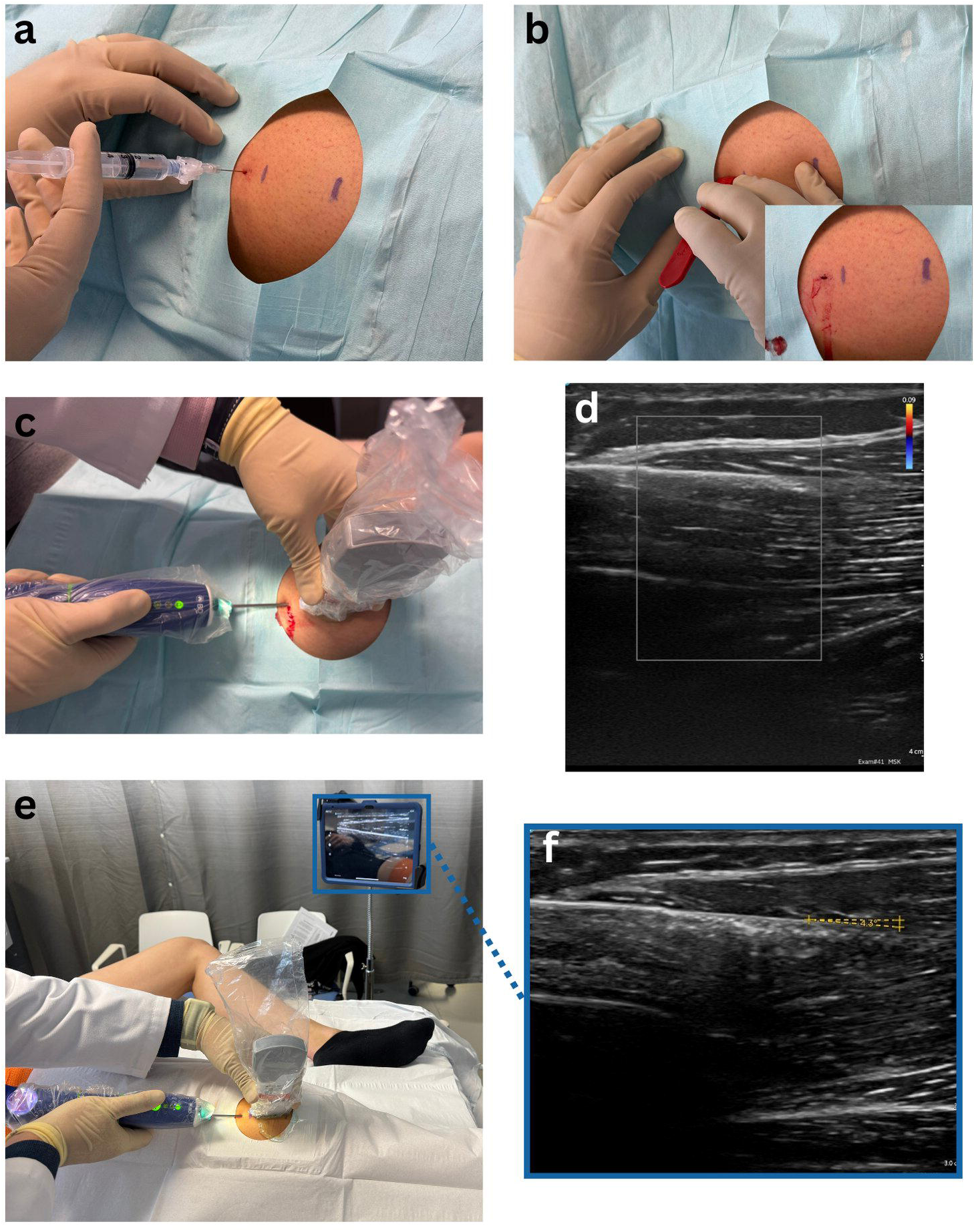
The muscle biopsy. A) Injection of local anesthetic (1% lidocaine) into sterilized biopsy site. B) Punch incision and resulting wound. C) Biopsy needle inserted into VL under sterile ultrasound guidance. D) Image from ultrasound application showing parallel alignment between muscle fibers and biopsy needle. E) Biopsy needle inserted into TA under sterile ultrasound guidance. F) Image from ultrasound application indicating a 4.3° angle between muscle fibers and biopsy needle.

#### Maintenance of the sterile field

The sterile operator and RA work together to provide sterile coverings of the non-disposable equipment such as the biopsy device and ultrasound probe. With the RA holding the base of either device, the sterile operator carefully covers the equipment with a sterile sleeve until they obtain a secure handle on the equipment and it becomes fully covered (Fig. 2c, *top*).

#### Biopsy procedure

After the deeper lidocaine has been delivered, a small incision (0.5cm) of the skin and subcutaneous tissue is made to facilitate biopsy needle entry (Fig. 3b). Through this incision, the sterile operator inserts the biopsy needle a single time into the VL. Ultrasound visualization, along with intermittent doppler use, are maintained through the procedure to ensure the needle is parallel to muscle fascicles and to avoid vasculature (Fig. 3c-f). 2-3 muscle samples are taken depending on participant tolerance, muscle size, and blood vessel location.

The needle is then removed, samples are transferred to the RA for processing, and the sterile operator dresses the wound. If the VL muscle biopsy procedure is tolerated well by the participant, they may elect to proceed with the muscle biopsy of the TA, following the same procedure phases as above.

#### Dressing the wound

After applying direct pressure with gauze to the incision, the wound is cleaned and dressed with Steri-Strip, an absorbent adhesive dressing, and a self-adherent compression wrap.

#### Specimen processing

2-3 specimens obtained from each biopsy are immediately weighed, blotted free from excess blood or extracellular debris, their dimensions measured, and they are allocated for further processing depending on desired histological, biomechanical, or biochemical analysis. Samples designated for biomechanical testing are permeabilized in dissecting solution with 0.5% w/v Brij 58 for 30 minutes with periodic agitation before transfer to storage solution for subsequent analysis [10]. For histology, specimens are pinned onto cork wrapped in parafilm to maintain orientation and frozen immediately in isopentane cooled by liquid nitrogen (−159°C). This step ensures optimal preservation of both structural and molecular features and prevents freezing artifacts that can compromise downstream analyses. Samples are stored at −80°C until further processing, which may include cryosectioning, histological staining, physiological testing, or other biochemical assays.

**Procedure** *(for the VL and TA)*:

**1.** Selection and marking of biopsy sites

**1.1.** Expose leg areas to be biopsied. Lay a disposable absorbent sheet under the exposed leg. Ensure the knee is extended, but relaxed and ankle is in neutral position throughout the entire procedure.

**1.1.1.** VL: expose preferred thigh from the groin crease.
**1.1.2.** TA: expose preferred ankle to knee
**1.2.** Ask participant to briefly tense/contract the muscle to outline the VL/TA. Mark biopsy site with a surgical marking pen. These recommended locations may be slightly altered depending on the presence and location of vasculature.

**1.2.1.** VL: Palpate hip region to locate the greater trochanter and locate the lateral femoral condyle. Draw or imagine a line between these two points and mark the procedure site approximately halfway between them, in the outer middle third of the thigh.
**1.2.2.** TA: Ask participant to internally rotate their leg approximately 30° towards the midline to activate the TA. Mark the procedure site as 6cm distal to the distal edge of the patella and 3cm lateral to the tibial crest.
**1.3.** Using a linear ultrasound probe in musculoskeletal mode, confirm the marked biopsy site is in alignment with the targeted muscle and there are no blood vessels in the area using the power doppler mode (Fig. 2e-f). Mark border of ultrasound device with surgical marking pen.
**1.4.** Mark the incision site 0.5 cm proximal to the proximal side of the ultrasound probe.
**2.** Prepare the skin

**2.1.** Wash hands with soap and maintain clean hands until sterile gloves are donned. The RA will prepare the biopsy tray by dropping all necessary sterile materials.
**2.2.** Clean the skin with a ChloraPrep applicator, starting from the incision site and working towards the periphery for 30 seconds in a 6 cm x 6 cm area.
**2.3.** Apply a sterile drape with a fenestration and adhesive aperture to expose the biopsy site while maintaining a sterile field.
**2.4.** Apply local anesthetic to the skin and underlying fascia at the incision site. Use a 25G x 1.5” needle to create a subcutaneous bleb of 1% lidocaine (Fig. 3a). After 60 seconds, infiltrate deeper into the subcutaneous tissue. Switch to longer 3.5” needle, if necessary, indicated by thick subcutaneous tissue above the muscle.

**2.4.1.** Aim to penetrate at least 1-3.5cm depending on the subcutaneous tissue thickness at the biopsy site (TA= 1cm, VL= 3-3.5cm). Maneuver the needle distally, into the region where muscle biopsy will take place.
**2.4.2.** Allow an additional 5 minutes for the anesthetic to take full action. Confirm anesthesia with the participant by probing skin gently.
**2.4.3.** Optionally, lidocaine delivery may be performed under ultrasound guidance if superficial vasculature or nerves are noted on the initial ultrasound scan.
**3.** Preparation of sterile instrumentation

**3.1.** Prepare ultrasound by having the RA place non-sterile ultrasound gel on the probe first before placing it in the sterile sleeve. The sterile operator will carefully slide the sleeve over the top of the ultrasound to grasp it while the RA holds the base of the non-sterile side. Once the sterile operator obtains a confirmed hold on the ultrasound, the RA may release the ultrasound which should be fully covered with the sterile sleeve at this point (Fig. 2c, *top right*).
**3.2.** After connecting the sanitized biopsy device to the needle, the RA should “prime” the collection receptacle by rotating, removing, and reinserting 5-8 times for easier access to samples. The RA should then cycle the device through the biopsy process one time to confirm proper function.
**3.3.** While the RA holds the base of the device steadily, the sterile operator covers the device with another sterile ultrasound sleeve, puncturing the sleeve with the biopsy needle and carefully rolling the sleeve down the length of the device until fully covered (Fig. 2c, *top left*). At this point, the sterile operator is the only person holding the device, similar to the process described in 3.1 above. Exercise great caution when putting sterile sleeve over exposed biopsy needle to avoid accidental needlestick injuries.
**4.** Biopsy procedure

**4.1.** Confirm participant feels no pain before proceeding.
**4.2.** With a size 11 scalpel, make a punch incision at the site of lidocaine injection, creating a 5 mm incision on the skin and down through to the underlying fascia (Fig. 3b).

**4.2.1.** Apply additional sterile gel to the now sterile ultrasound probe.
**4.3.** Using the sterile ultrasound probe and gel, with doppler effect on, guide the biopsy needle into the muscle distally and along the long axes of the femur/tibia, ensuring the needle is as parallel to the muscle fascicles as possible (Fig. 3c-f). Typically, initial angle of entry is approximately 30 degrees relative to the skin, but will be adjusted in real time to be parallel with fascicles

**4.3.1.** Retract the needle on the biopsy device before entry into tissue using the designated button
**4.3.2.** One the needle is aligned for collection, eject the needle using the same button as used for retraction.
**4.3.3.** Confirm needle placement again on ultrasound and using doppler, then turn off doppler mode and press the sample button for sample collection.
**4.4.** Repeat sample collection without withdrawing the needle or removing the collection container to maintain sterile field. Multiple samples may be collected in the same container.

**4.4.1.** For additional samples, either advance the needle 2-3 cm distally along the muscle fibers or redirect the needle in the superficial to deep plane. Maintain doppler mode when advancing or redirecting to visualize vessels.
**4.5.** When sufficient samples are collected, withdraw the needle.
**4.6.** With the operator holding the sterile biopsy device, maneuver the device out of the sterile sleeve to expose the unsterile collection receptacle. The RA then removes the tissue collection container, removes the specimen, and returns the container to the biopsy driver. The device operator then slides the sterile sleeve back onto the driver. This allows samples to be retrieved without breaking the sterile field.
**4.7.** The first biopsy site is closed according to the post procedure in step 6 before a second biopsy is collected in any other site, repeating steps 2-4.
**5.** Dressing the wound

**5.1.** Immediately after needle removal, apply direct pressure to the wound with gauze for up to 2-3 minutes. Close the wound with two Steri-Strips by placing them in an “X” formation.
**5.2.** Use a pre-filled saline syringe flush as needed to clean the skin around the biopsy site.
**5.3.** Place a sterile absorbent adhesive dressing over the Steri-Strips and wrap a self-adherent compression bandage around biopsy site and secure with tape.
**5.4.** Ask the participant to rate their immediate post-procedure pain on the Visual Assessment Sale (VAS) from 1-10 and document their answer.
**6.** Sample processing/storage (for the non-sterile RA assistant)

**6.1.** Immediately after removing the samples from the collection container, blot off excess blood on gauze. If samples are exceptionally bloody, place them in a 6 well plate containing 1x PBS. Move the samples through a serial dilution until the PBS no longer turns red (Fig. 4a). Blot off excess PBS.
**6.2.** Weigh samples on a portable scale with 0.001g sensitivity and draft shield for accuracy and record weight.
**6.3.** Prepare sample for long term storage as appropriate for the intended analysis:
**6.4.** Flash freezing and histological analysis: use minute stainless steel pins to secure samples to cork wrapped in parafilm. Pin samples with *gentle* tension to retain fiber length during the freezing process (Fig. 4b). Place the cork in a 50ml conical tube, stored in a pre-cooled rack.

**6.4.1.** Once the procedure is complete, samples can be flash frozen. Pre-cool isopentane in liquid nitrogen for 60 seconds. Place samples into the cooled isopentane for approximately 10 seconds and store samples at −80°C until further use (Fig. 4c-d).
**6.5.** Respirometry: blot samples free of excess blood and cut an ∼30mg of tissue to place immediately in ice-cold BIOPS solution, maintain at 4°C for further respirometry processing as soon as possible, typically within 1-2 hours.
**6.6.** RNA extraction: specimen can be placed in RNAlater. These tubes should be stored at 4°C for 24-72 hours before long term storage in −80°C.
**6.7.** Single fiber biomechanics: Remove sample from biopsy device and place in 5 ml of chilled dissecting solution. Transfer sample to 700 µl of dissecting solution with Brij over ice for 30 minutes, gently shaking every 5 minutes. Transfer samples back to original 5 ml dissecting solution for 5 minutes, shaking every minute, to wash off excess Brij 58. Transfer each sample into 1 ml of chilled storage solution for long-term storage at −20°C.
**7.** Preparing specimens for histology

**7.1.** Samples should be removed from storage at −80°C and placed in a cryostat set to −20°C for 45min-1hr to slowly come to temperature. All necessary tools should be placed in the cryostat.
**7.2.** Using a single edge razor blade, cut directly perpendicular to the muscle fibers to obtain an approximately 0.5cm long core (Fig. 5a-I). Let this sample acclimatize to the cryostat temperature for another 20 minutes.
**7.3.** Prepare a 0.5cm^2^ cryomold and fill with frozen section media (Leica Biosystems #3801480)

**7.3.1.** Let cool in cryostat for approximately 45 seconds to slightly harden. This will minimize freezing artifact.
**7.3.2.** Using forceps, quickly place the specimen into the cryomold so that the muscle fibers are perpendicular and flush to the base of the mold (Fig 5a-II).
**7.3.3.** Using long, 300mm specimen forceps, quickly invert the mold into isopentane cooled in liquid nitrogen for 10 seconds (Fig 5a-III) and return to cryostat. Let this flash-frozen mold acclimate to the cryostat temperature for 20 minutes.
**7.4.** Remove the specimen from the mold. On the chuck, add a ring of frozen section media and quickly and gently place the specimen onto the chuck with forceps. The side of the specimen that was closest to the base of the mold should have a flat surface and now be facing the technician, superficial to the chuck, as seen in Fig. 5a-IV.
**7.5.** Section the specimen at 8-10µm thick. Use fine tip brushes to orient or flatten sections before placing them on labeled microscope slides (Fisher Scientific #12-544-2).
**7.6.** Perform a quality check under a brightfield microscope to confirm correct orientation and proper cross sections of the muscle specimen.
**7.7.** Store slides if needed long term at −80°C or immediately proceed to staining protocol.
**7.8.** Stain resulting slides with histochemistry (Fig. 5b, c) or immunohistochemistry (Fig. 5d) as desired.

**Figure 4:**
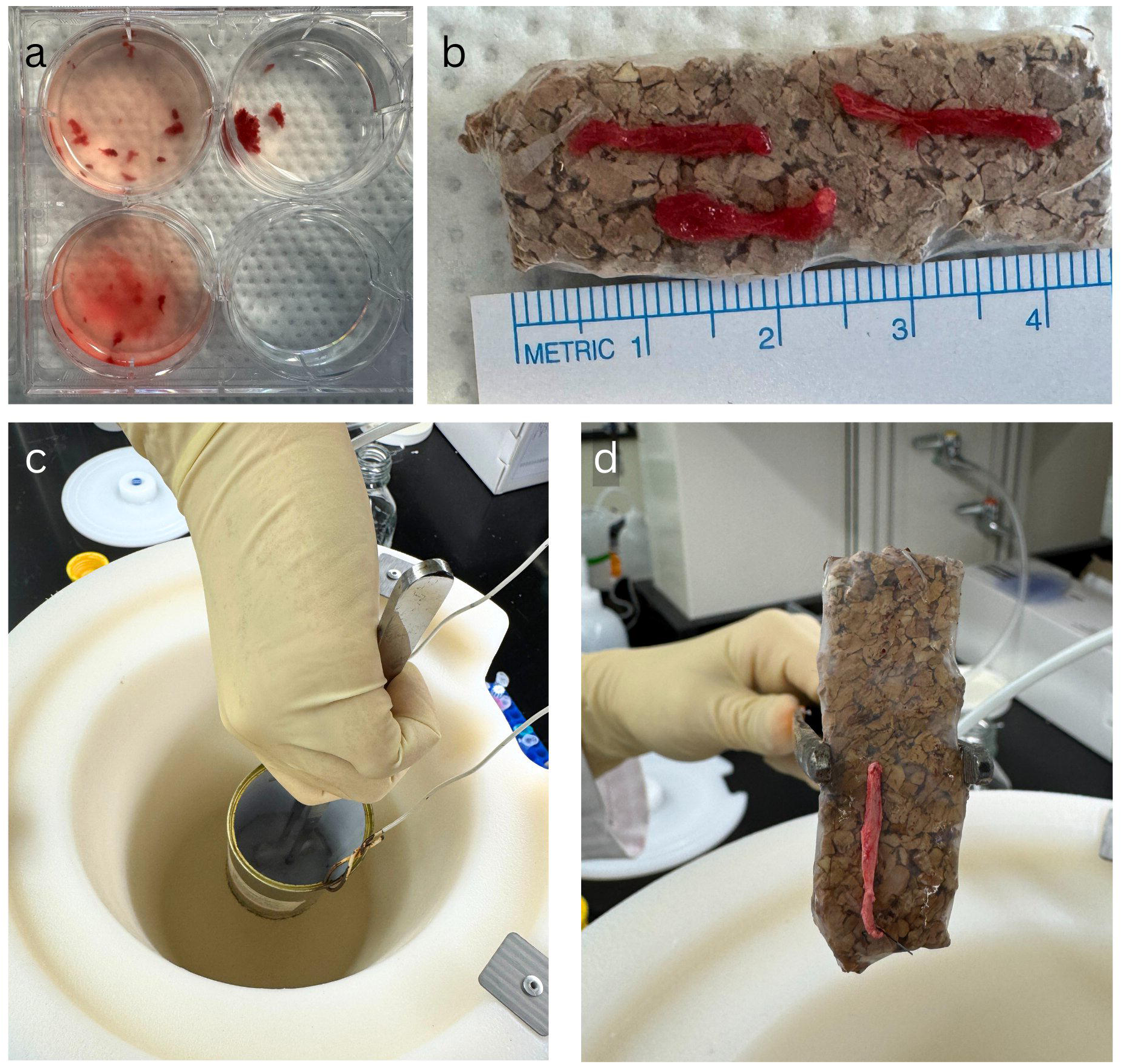
Specimen processing for long-term storage. A) Successive rinsing technique to clear debris/blood from specimen. B) Specimen from the VL after rinsing on parafilm-wrapped cork for freezing. C) Freezing specimen in isopentane cooled in liquid nitrogen. D) Specimen after freezing, muscle fiber shape is maintained with slight tension from minute pins at either end.

**Figure 5:**
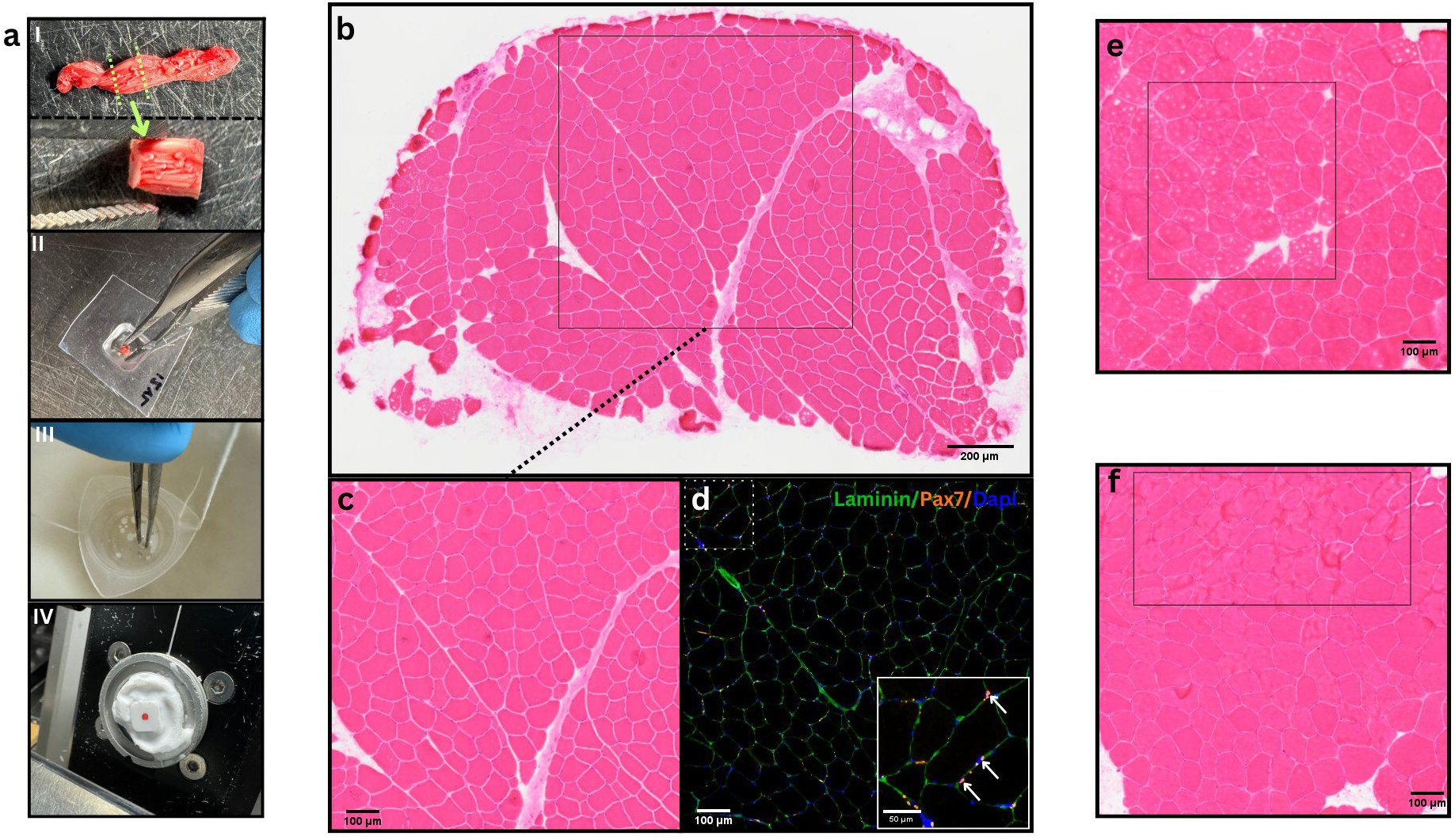
Histology. A) Process of preparing samples for histology. I) Frozen specimen with green lines indicating region to be cut directly perpendicular to visible fibers. II) Adding sample to OCT in a 0.5cm2 mold III) Flash freezing the sample & mold in isopentane cooled in liquid nitrogen. IV) resulting specimen frozen in OCT and mounted on cryostat chuck. B) representative image of whole section H&E of VL sample. C) 20x resolution of VL from A. D) Laminin/Pax7/DAPI immunofluorescence of serial VL sections from A. Arrows in inset image indicating Pax7+ cells. E) H&E artifact resulting from improper duration of freezing specimen. F) H&E Artifact resulting from folding of specimen during cryosectioning.

### Time Required

The entire procedure to obtain both VL and TA muscle biopsies requires approximately one hour. Setting up the procedure space and the visualization/marking of the biopsy sites with ultrasound guidance requires ∼15 minutes. Once the sterile field is established, the biopsy requires ∼15 minutes, that includes administration of local anesthetic, creating the incision, and obtaining muscle specimen. Duration of the biopsy procedure, determined from when the needle is first inserted, samples are collected, and needle is removed, is less than ∼5 minutes. Dressing the wound and monitoring for any immediate complications from the procedure requires approximately 15 minutes.

### Post-biopsy suggestions for care

After completion of the procedure, muscle stiffness is commonly experienced by the participant and may be relieved by gentle exercise or movement (e.g., walking). As needed, simple analgesia such as acetaminophen is recommended for pain or soreness as well as applying ice to the site in a 20 minutes on/20 minutes off cycle. It is recommended participants avoid taking Aspirin, ibuprofen (Advil), or other NSAIDs as they may increase the risk of bleeding.

To increase the likelihood of participant compliance with these recommendations, it is imperative they receive written directions the care of their wound and are educated on signs of infection or complications, such as excessive swelling, tenderness, or redness. The participant should remove the self-adherent compressive wrap 6-8 hours after completion of the procedure. They may remove the absorbent adhesive dressing after 24-48 hours and the adhesive wound closures/Steri-Strips should be left to fall off on their own within 1 week or may be removed after 10 days. Subsequently, a Band-Aid or other adhesive bandage may be used to cover the site if the participant desires but this is typically not necessary.

The recovery period is expected to be 3-4 days with the greatest amount of discomfort in the 6-8 hours immediately following the biopsy procedure. Participants should be instructed to avoid strenuous physical activity (e.g., running, lifting heavy objects, sports activity, biking, etc.) for 48 hours and should return to normal activity levels as they feel comfortable. The participant should avoid submersion in water (e.g., swimming in a pool/lake, taking a bath) for 72 except for showering in which case the participants should wrap cling film (or Saran/plastic wrap) around the site to keep the dressing dry.

### Troubleshooting

During the development of this protocol, several issues arose that may be important and are reported here for transparency and reproducibility.

#### Pain with needle insertion

Pain is usually reported when the biopsy needle crosses the fascial plane and enters muscle tissue. If discomfort is too high, the operator may elect to remove the needle and administer more local anesthetic. In some cases, a higher depth needle is indicated by the appearance of subcutaneous tissue thicker than 3.5 cm upon ultrasound visualization.

#### Adhesive allergies/alternatives

Allergies to adhesives present a challenge when deciding how to properly close and dress the biopsy wound but are not a contraindication for this procedure. An alternative closure method we recommend is the use of 5-0 absorbable sutures for closure and dressing the wound with 2-3 absorbent gauze pads held in place with a self-adherent compressive wrap. The compressive wrap and gauze should then be exchanged every 8-12 hours for the first 48 hours. This allows the wound to heal properly without scarring or irritation for most people with a sensitivity to adhesives. It is important to note that, with this modification; the physician should take care to inject additional local anesthetic into the superficial tissue to alleviate any pain that may be experienced from closing the wound.

#### Clearing debris/blood from tissue

Excess blood or coagulation of blood may interfere with specimen processing and tissue quality for histological or biochemical analysis. Specimens should first be blotted with slightly damp gauze. If this is insufficient to debride the sample, successive rinsing with 1x PBS is recommended (Fig.4a). This includes moving specimens through multiple successive washes in fresh saline to gradually remove excess blood. Saline should then be removed by blotting specimen on damp gauze.

#### No specimen visible in chamber

If the needle is retracted but no specimen is visible in the collection receptacle, this indicates an error with the vacuum system. If the cutting window of the needle is between muscle and subcutaneous tissue (at the fascia), the device may be unable to achieve adequate suction and therefore no sample is retrieved. Alternatively, after multiple biopsies, blood or tissue remaining in the needle may prevent successful specimen retrieval.

Specimens may be trapped in the cannula and may be retrieved by cycling through ejection and retraction of the needle once the device is removed from the participant. If this technique fails, it may be necessary to change the needle before attempting any further biopsies or reinserting the needle.

#### Specimen being obtained in fragments

This may be a result of inadequate suction if the cutting window is across the fascia betwen subcutaneous and muscle tissue. Alternatively, shearing force of the needle retraction may result in the fragmentation of the specimen. This is more common for the second and third sample collections within the same incision and may result from taking a sampling from the same physical location repeatedly, retrieving damaged tissue that is more easily fragmented. In either case, we suggest redirecting or advancing the needle as outlined above so the specimen is retrieved from an adjacent section of muscle and the cutting window no longer crosses the fascial plane.

#### Histological Artifact

Freezing artifacts may appear on histological stains or sections of muscle samples if the specimens are place in isopentane for either insufficient or extended durations (Fig 5e). No longer than 10 seconds is recommended for this freezing process. Artifact may also occur from excess saline during sample processing. To avoid these artifacts, properly blot the sample on damp gauze before freezing so that it remains moist without being oversaturated. Folding of the specimen may appear (Fig. 5f) during the sectioning process. This is avoided by carefully flattening each section with a fine tip brush as it adheres to the microscope slide.

#### Oblique Cross Sections

Oblique cross sections may be observed during initial cryosectioning of samples after performing the quality check under a brightfield microscope. Optimal muscle cross sections should have a uniform polygonal shape with no obvious major and minor axes. Oblong or oval muscle fibers indicate improper orientation of the specimen relative to the cutting blade that should be corrected by slight adjustments to the angle of the specimen chuck, repeated sectioning and brightfield visualization. This process may need to be repeated before optimal orientation is achieved.

### Methods for Further Analysis

All data were analyzed with GraphPad Prism 10. Significance was set as p<0.05 for all analyses. Results are reported as mean±SD unless otherwise noted. Measurement of gross sample dimensions was performed in ImageJ [23]. Analysis of fiber cross sectional area (fCSA) was performed using MuscleJ [24].

#### Histology

After sectioning muscle samples as described above, histological analysis was performed using the traditional hematoxylin and eosin stain as well as immunofluorescence (Fig. 5d). Sections were stained for laminin myofiber border protein (Sigma-Aldrich #L9393, Invitrogen #A10680) and 10,000µm^2^ 20x magnification Images from each sample were used to quantify fiber number and fCSA (Fig. 5d). These sections were also immunofluorescently stained for the muscle satellite cell marker, Pax7 (DSHB, University of Iowa, PAX7), as previously described [25]. Pax7+ cells, as defined by the co-localization of Pax7 with DAPI within fiber boundaries (Invitrogen #62248), were manually quantified and confirmed with automated quantification using MuscleJ [24].

*RT-PCR:* A portion of the muscle specimens was placed in RNALater (Invitrogen #AM7021) and stored overnight at 4°C before long term storage at −80°C. RNA was extracted (Qiagen RNeasy Mini Kit, #74104), converted to cDNA (Bio-Rad iScript cDNA Synthesis Kit, #1708890), and RT-PCR was conducted for genes indicative of normal muscle function [26,27]: myosin heavy chain 1 (*MHC1*), myosin heavy chain beta (*MHC7*), troponin (*TNNT3)*, as well as genes relating to myogenesis: myogenin (*MYOG)* and myogenic differentiation factor 1 (*MYOD1*). Beta-actin (*ACTB*) was the reference gene and results are reported as 1-dCq values.

#### Collagen content

Collagen content was determined using the hydroxyproline assay as previously reported [28,29]. Briefly, 5-10mg of each biopsy sample was hydrolyzed in concentrated acid for 24 hours before being plated and evaporated over an additional 24 hours. Samples were incubated with a Chloramine-T solution at room temperature for 20 minutes before being incubated with a p-dimethylaminobenzaldehyde solution for 30 minutes at 60°C. Colorimetric absorbance was read at 550nm and plotted against a standard curve.

Hydroxyproline concentration was multiplied by the collagen constant (7.46) to give the final collagen concentration in each sample.

#### Mitochondrial Respirometry

High-resolution mitochondrial respirometry was performed using the Oroboros O2k system (Oroboros Instruments, Innsbruck, Austria) following established protocols [30,31]. Briefly, within an hour of biopsy collection, muscle tissue was mechanically separated in ice-cold BIOPS under a dissecting microscope, chemically permeabilized with saponin (25µg/ml, 20 mins), and washed in mitochondrial respiration media (MiR05) (10 mins) on a rocker at 4°C. Samples were gently blotted on a Whatman filter paper, weighed, and immediately transferred to calibrated Oroboros O2k chambers containing MiR05.

Respirometry experiments were conducted in duplicate at 37 °C under hyperoxygenated conditions. Sequential substrate-uncoupler-inhibitor titration (SUIT) protocol (pyruvate, malate, ADP, glutamate, succinate, uncoupler, rotenone, antimycin a) were used to assess complex I- and II-supported oxidative phosphorylation, maximal electron transport system (ETS) capacity, and residual non-mitochondrial respiration. Cytochrome c addition was used to verify outer mitochondrial membrane integrity, and replicates with >10–15% respiration increase were excluded. Oxygen flux was normalized to tissue wet weight and reported as pmol O ·s ¹·mg ¹.

#### Single Fiber Mechanics

Active and passive biomechanical properties were determined on permeabilized single fibers isolated from muscle fiber bundles as previously described [10,32–33]. Briefly, fibers were teased from bundles in a relaxing solution (pCa 9.0; [10]) and mounted in the first bath of the permeabilized fiber testing system (1400A; Aurora Scientific) using 10-0 silk suture. Sarcomere length was set to 2.7 µm [10] using an inverted light microscope with a Fourier based deconvolution. Fiber diameter was measured in three places along the length of the fiber and fiber cross-sectional area was calculated using the average diameter and assuming a cylindrical cross-section.

Following these measurements, fibers were rapidly transferred to a bath with a weakly buffered pre-activating solution, followed by a high Ca^2+^ activating solution (pCa 4.5; [10]). Force development was recorded from transfer to activating solution (up arrow, Fig. 9a) through return to relaxing solution (down arrow, Fig. 9a). This process was repeated three times and average peak active force calculated. After active testing, fibers were equilibrated in relaxing solution for 5 minutes prior to initiation of a passive incremental stress relaxation test. For this test, fibers were stretched in 200 µm increments at 4 mm/sec with 4-minute holds for 6 stretches or until failure, with simultaneous force and sarcomere length recording (Fig. 9b). All forces were normalized to fiber cross-sectional area to obtain stresses. Peak and relaxed passive stresses were fit to sarcomere strain to determine peak and relaxed passive modulus for each fiber (exemplar values shown with filled circles in Fig. 9b).

## Results

The muscle biopsy procedure was well tolerated by all 19 participants. Average VAS pain rating immediately after the procedure was 1.5±1 (range: 0-4, Fig. 6a). 24 hours after completion, average VAS rating was 1.7±1 (range: 0-4). Adverse reactions or observations were recorded and reported in Figure 6b. Dizziness, nausea, or faintness during or immediately after the procedure occurred in 4 of the participants and dissipated after 15 minutes. Short-term reactions (onset within 6 hours of the procedure) occurred in 2 participants. Bleeding through the compression wrap was reported in one instance of the TA which was controlled with additional compression over the next 8 hours. One participant experienced numbness and weakness in the ankle/foot after the TA biopsy and was monitored closely by the physician. This weakness subsided within 20 minutes after completion of the procedure. Long term reactions (onset >24 hours after procedure completion) occurred in one participant who noticed a tight “muscle cramp” like symptom in the calf region of the leg 72 hours after the TA was biopsied in that leg. Although this was likely an unrelated incident (since no calf muscles were biopsied), we report it here for transparency.

**Figure 6:**
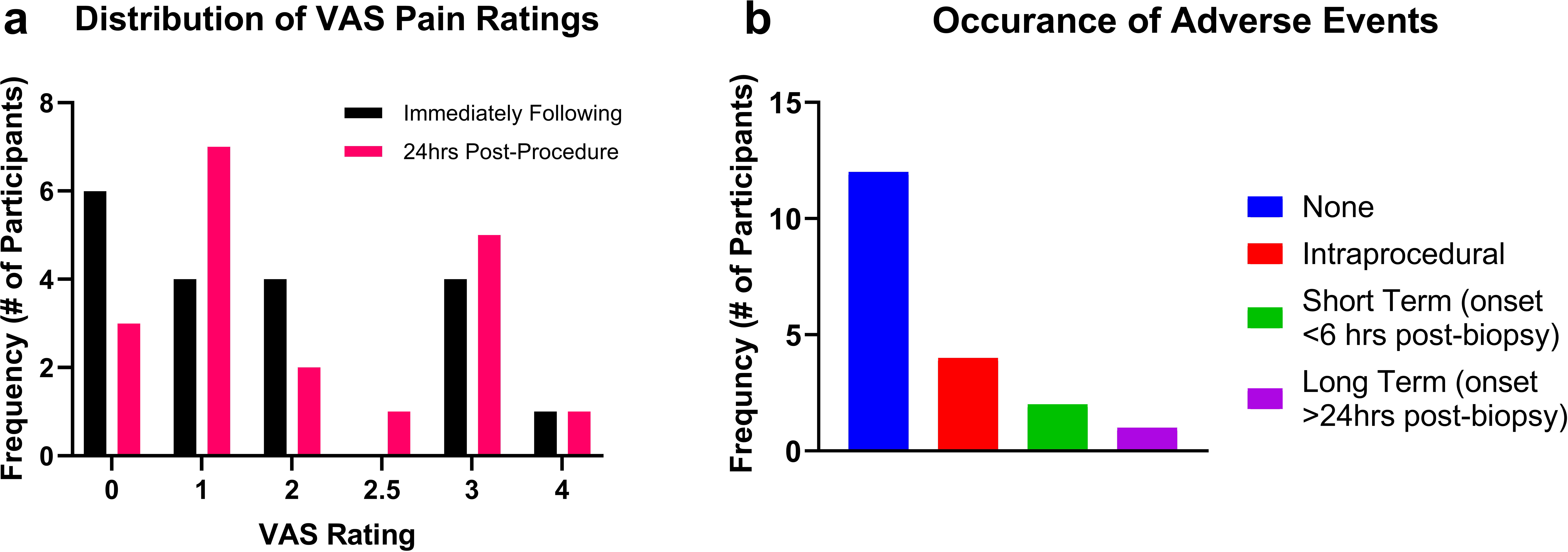
Patient outcomes. A) Distribution of pain ratings using the Visual Analog Scale (VAS) immediately after completion of the biopsy procedure and 24hrs later. B) Occurrence of adverse reactions. Intraprocedural includes faintness/nausea/dizziness that resolved before completion of the procedure. Short term reactions include issues that had an onset within 6 hours of procedure completion and were resolved within 24 hours. Long term reactions included those with an onset >24 hours following procedure completion.

Of the 19 participants enrolled, all 19 completed a muscle biopsy from their VL in which 2-3 muscle samples were obtained through a single needle insertion. 16 of these participants also completed the TA biopsy. 2 participants were not biopsied at the TA due to scheduling constraints; 1 participant elected to not proceed due to an adverse reaction to the VL biopsy. TA biopsy specimens from 2 participants were not obtained or included in analysis due to unsuccessful specimen retrieval, likely due to blood or tissue in the device cannula interfering with the internal vacuum system as described above. In total, 53 VL samples and 41 TA samples were collected from 19 participants.

The mass and gross dimensions of VL and TA samples were compared by unpaired t-tests. There was no significant difference in average length of VL (15.83±8mm) compared to TA (15.07±7mm) samples (p>0.6; Fig. 7a) even though the actual fascicle length of the two muscles is quite different [22,34]. This may reflect an upper limit of biopsy length determined by the 20mm length of the sample collection window (Fig. 2b). There was no significant difference in average width of VL (2.91±0.6mm) compared to TA (3.08±0.9mm) samples (p>0.3; Fig. 7b).

**Figure 7:**
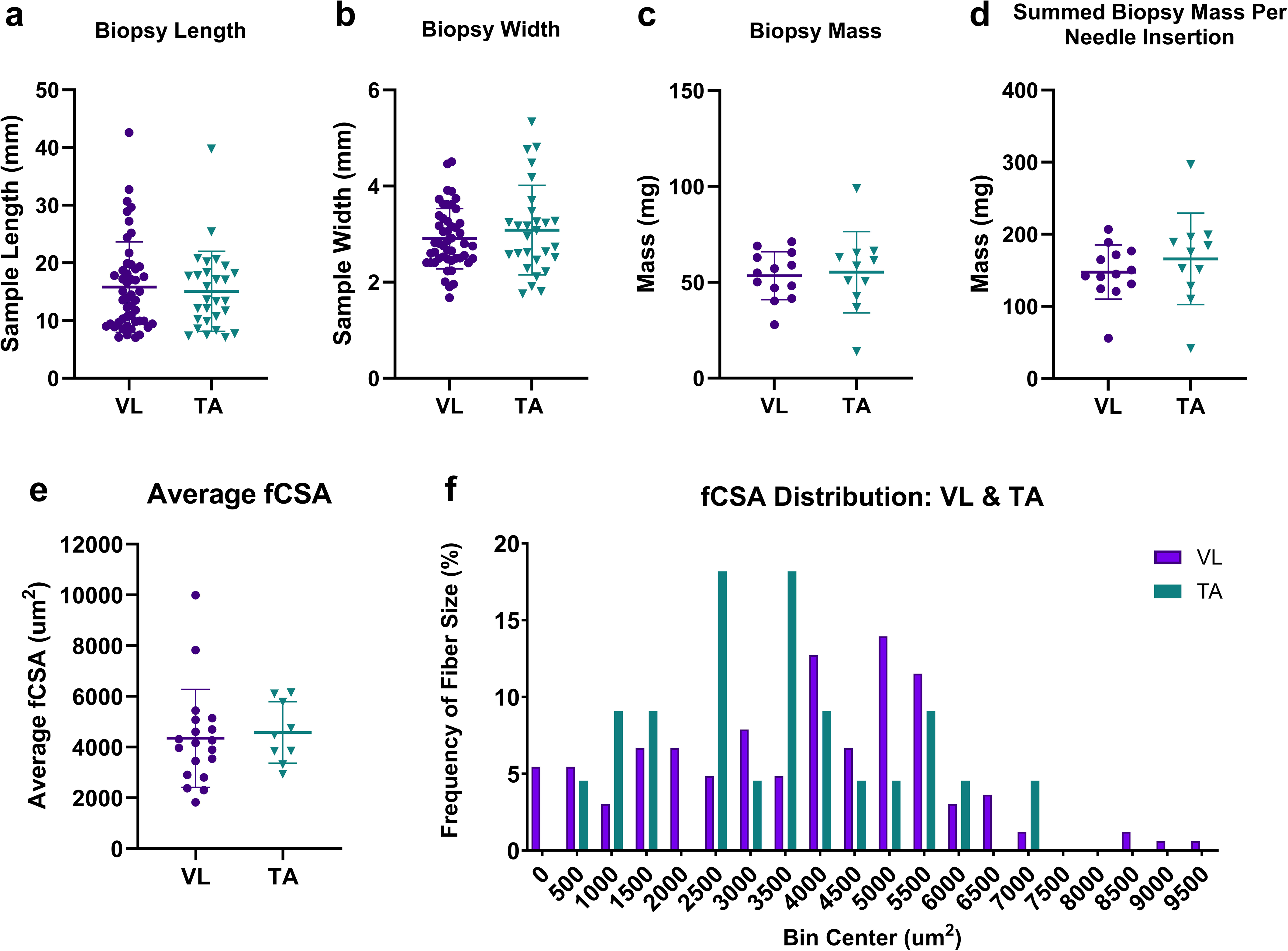
Gross dimensions of VL and TA biopsies. A) Average biopsy length. B) Average biopsy width. C) Average biopsy mass of a single sample. D) Summed biopsy mass of three muscle samples from a single needle insertion. E) Average fCSA. F) Frequency distribution of fCSA (µm^2^) in VL and TA samples for the representative participant in Figure 5. Data reported as mean±SD. Statistical analysis conducted for A-E unpaired t-tests. α was set to 0.05.

Average mass of VL samples was 53.4±12mg and average TA mass was 55.3±21mg (p>0.7; Fig. 7c). Multiple samples are taken from each muscle with a single needle insertion and, when summed together, the average biopsy yield from the VL was 148±38mg and 166±64mg from the TA (p>0.4; Fig. 7d). Although some studies [35,36] using the Bergström technique have reported yields of 100-200mg, the success of this technique is highly user dependent or relies on wall suction which may not be available in all settings.

To assess sample quality for histological analysis, we performed traditional Hematoxylin & Eosin stains on 10µm thick sections which confirmed the proper orientation of the muscle cross section, indicated by their neatly organized polygonal shape (Fig. 5b-c; [2]). Average number of VL and TA muscle fibers quantified were 221±86 and 202±72 respectively (p>0.5). Average fCSA of VL and TA muscles was 4,347±1931 µm^2^ and 4,576±1,208 µm^2^, respectively (p>0.7; Fig. 7e). A histogram depicting the frequency distribution of fiber sizes in the VL and TA of the representative participant from Figure 5b-d is shown in Figure 7f. VL and TA fCSA for this participant was 3892±2370 µm^2^ and 3850±2888 µm^2^, respectively. These data, in conjunction with the mass and gross dimensions of these samples, confirm that the technique described here consistently yields muscle samples of high enough quality to yield morphometric data.

Portions of the biopsy were allocated for further histological and biochemical analysis to demonstrate the quality of sample obtained and potential applications for future research in muscular disease. Muscle satellite cells are required for the repair/regeneration of healthy muscle tissue [37,38]. In the VL and TA, the number of Pax7+ cells per 100 fibers were 06.06±6.7 and 9.09±7.3, respectively (p>0.3; Fig. 8a). The presence of Pax7+ cells in our muscle samples further indicates proper specimen processing in this protocol and has implications for further comparison in future studies of muscle disease. RT-PCR 1-dCq values of these healthy controls show appropriate expression of genes commonly associated with normal muscle function and myogenesis (Fig. 8b). Fibrosis is a common feature of muscular disease [2,3,39], making intramuscular collagen content an important area of investigation. Average collagen content of VL and TA samples was 3.85±3.1µg collagen/mg tissue and 4.74±2.9µg collagen/mg tissue, respectively (p>0.4; Fig. 8c). Mitochondrial respiration was evaluated in fresh, permeabilized samples (VL: n=7, TA: n=8) to quantify maximal oxidative phosphorylation and electron transport chain capacity. As expected, two-way mixed effects ANOVA showed a main effect of substrate (p<0.0001, Fig. 8d) but no effect of muscle tested. Maximal electron transport chain capacity was 99±30 pmol/s/mg and 122±43 pmol/s/mg for the VL and TA, respectively, slightly higher than our previous results with open biopsies in older individuals (∼72 years of age).

**Figure 8:**
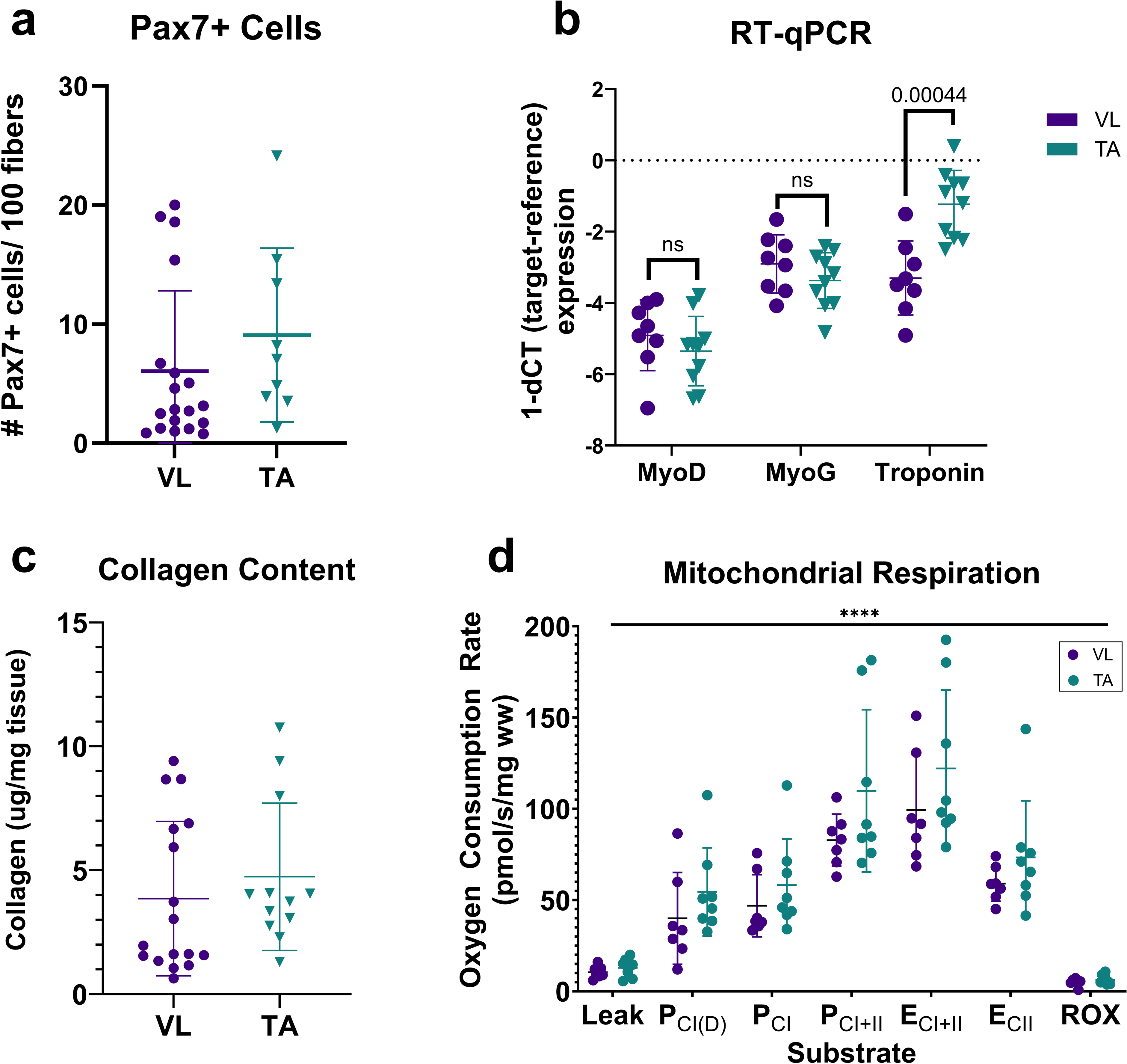
Additional applications. A) Pax7+ satellite cells per 100 fibers in VL and TA samples. B) RT-PCR gene expression results, normalized to actin-beta, reported as dCq values. C) Collagen content of VL and TA samples D). Mitochondrial respiration in fresh, permeabilized VL and TA samples. Sequential protocol of Pyruvate, Malate (Leak), ADP (PCId), Glutamate (PCI), Succinate (PCI+II), FCCP (ECI+II), Rotenone (ECII), Antimycin A (ROX) was used to test maximal oxidative phosphorylation and electron transport chain capacity. (2-way mixed effects ANOVA of substrate x muscle showed substrate effect p<0.0001). Data reported as mean±SD. A-C were analyzed with unpaired t-tests, α was set to 0.05.

Active and passive fiber biomechanical properties were measured from TA samples (Fig. 9). Passive elastic modulus based on peak and stress-relaxed force was 239 and 78.6 kPa respectively (Fig. 9b). Average active stress was187 kPa (Fig. 9a), similar to results previously reported for experiments performed at this temperature [10], confirming the feasibility of this method to produce intact fiber bundles for testing.

**Figure 9:**
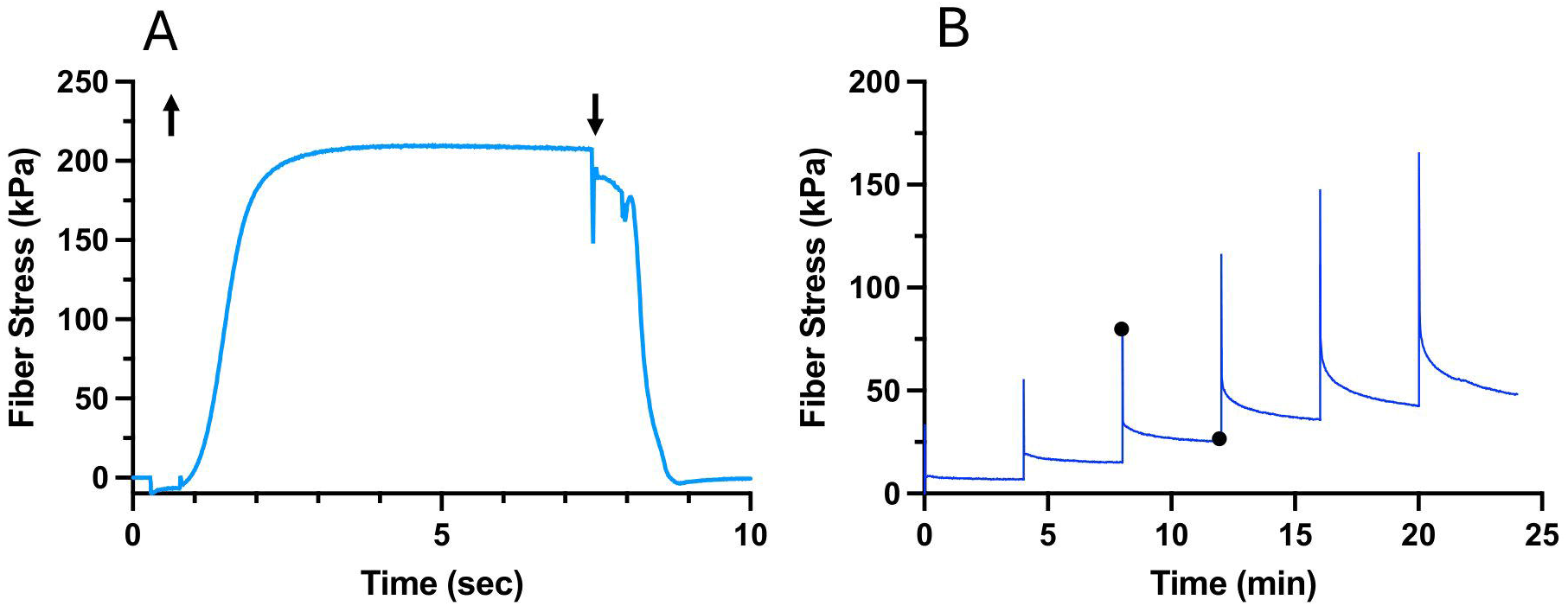
Fiber mechanics. A) Continuous display of single muscle fiber stress during chemical activation and relaxation. Fiber is set to a resting sarcomere length of 2.7 µm and transferred from relaxing solution to activating solution (up arrow) and force allowed to develop and stabilize. Then, the fiber is transferred to relaxing solution (down arrow) and force returns to baseline. B) Continuous display of single muscle fiber stress during stepwise elongation as described in methods. Immediately after stretch, force increases and then stress-relaxes over the subsequent period (4-minutes). These data yield single fiber modulus of peak and stress-relaxed values (filled circles) for fiber modulus, thus characterizing its passive viscoelastic properties.

## Discussion

Here we describe a systematic approach to muscle biopsy collection in the vastus lateralis (VL) and tibialis anterior (TA) which, combined with ultrasound guidance, yields samples of sufficient quality and quantity for simultaneous histological, mechanical, biochemical, and metabolic analysis, as demonstrated by the high-quality histological stains, fCSA quantification, appropriate active and passive physiology testing, as well as multiple successful methods of downstream analysis. In comparing results obtained from VL and TA, we observed no significant differences between the two muscles with regard to sample size, mass, muscle fCSA, Pax7+ expression, and collagen content. It is our intention to apply the protocol described here to further investigate the nature of human muscular disease.

This approach overcomes several limitations of traditional muscle biopsy techniques, as outlined in Table 2. Open biopsies consistently provide large samples that may preserve fiber orientation but often are dependent on access to an operating room, carry higher risks of hematoma and scarring, and require sutures or general anesthesia. Bergström and suction-assisted needle techniques, while less invasive, may yield inconsistent sample sizes and lack real-time visualization, increasing the risk of sampling error or inadvertent neurovascular injury.

**Table 2:**
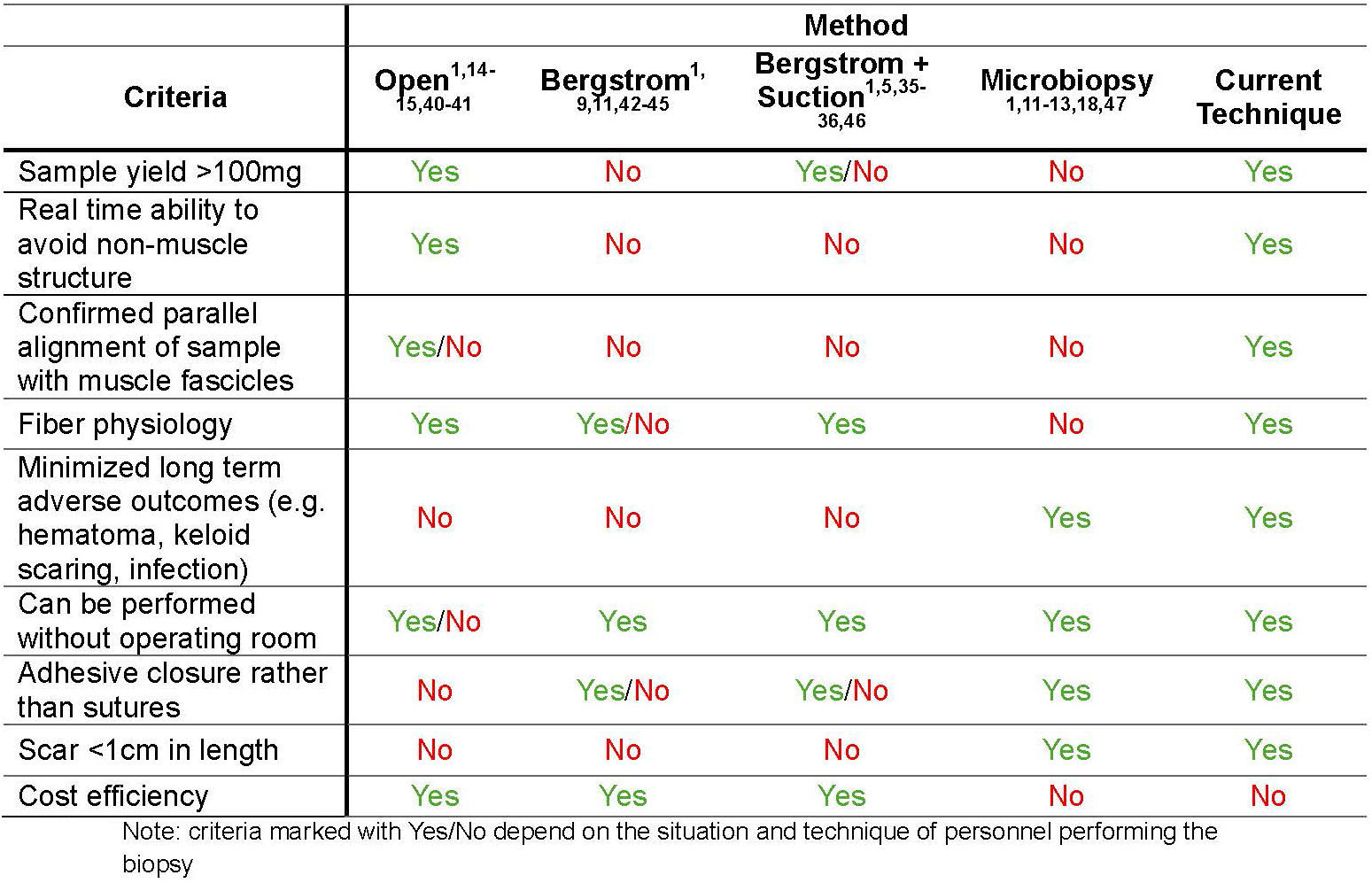
Comparison of muscle biopsy methods.

Microbiopsies minimize invasiveness but frequently produce insufficient tissue for comprehensive histological, physiological and biochemical analyses. In contrast, our protocol combines real-time ultrasound guidance with a vacuum-assisted SIMS device to optimize sample orientation, avoid non-muscle structures, and obtain multiple high-quality samples from a single insertion. This strategy enables robust downstream analyses while reducing procedural complications. This approach represents a practical balance of diagnostic capability, patient safety, and procedural efficiency.

A current limitation of our method is the limited selection of muscles biopsied. However, VL and TA muscles are some of the most highly characterized muscles in terms of intramuscular variability and implications in numerous muscular diseases [2,19,22]. Therefore, it seemed essential to first develop this protocol on the muscles most commonly assessed in clinical and research settings. Applying this technique to other muscles should be entirely feasible but further validation is needed to optimize this protocol for other muscles. This can be easily done by leveraging the ultrasound component of this method for future planning. Additionally, the participants reported here consisted primarily of younger individuals who may tolerate this procedure better than an individual with an underlying disease or in an older population.

However, as our primary goal is to describe this muscle biopsy technique to increase reproducibility in the field, we hope to fuel further research into these other patient populations including aging and chronic disease conditions.

## Acknowledgments

The authors thank all the participants who volunteered for this research. The authors also thank BioRender for aid in the creation of Figure 1. This work was supported by a Foundational grant from the Catalyst Grant Program of the Shirley Ryan AbilityLab. IR is supported by a career development award through the National Institute of Arthritis and Musculoskeletal and Skin Diseases (K08 AR081391). This work was also supported in part by Research Career Scientist Award Number IK6 RX003351 from the United States (U.S.) Department of Veterans Affairs Rehabilitation R&D (Rehab RD) Service.

## Author contributions

A.W., G. Meza, J.F., S.D., and G. Meyer were involved in data collection and editing of this manuscript. R.L was involved in the development and editing of this manuscript. A.B. and I.R were involved in the development, data collection, writing, and editing of this manuscript.

## Ethics Declaration

This human subjects study was approved by the Northwestern University Institutional Review Board approved on 6/14/2024 (IRB ID: STU00221110). All methods were performed in accordance with the relevant guidelines and regulations. All participants gave written and informed consent.

## Data availability

Raw data obtained using this protocol are available from the corresponding author upon request.

## Additional information

Correspondence and requests for materials should be addressed to I. R.

